# Vaginal microbiome is associated with endometrial cancer grade and histology

**DOI:** 10.1101/2022.01.31.22270189

**Authors:** Hesamedin Hakimjavadi, Sophia George, Michael Taub, Leah Dodds, Alex Sanchez-Covarrubias, Marilyn Huang, Matthew Pearson, Brian Slomovitz, Erin Kobetz, Raad Z. Gharaibeh, Ramlogan Sowamber, Andre Pinto, Srikar Chamala, Matthew Schlumbrecht

**Author notes:** Co-Senior & Corresponding Authors <u>Corresponding Authors:</u> Matthew Schlumbrecht, MD, MPH, Professor of Gynecologic Oncology, Sylvester Comprehensive Cancer Center, University of Miami, 1121 NW 14^th^ St, Suite 345C, Miami, FL 33136, Srikar Chamala, Ph.D., Clinical Assistant Professor, Director of Biomedical Informatics, Dept. of Pathology, Immunology and Laboratory Medicine, Dept. of Health Outcomes and Policy University of Florida, P.O. Box 100275, Gainesville, FL 32610. Contributed equally (Hesamedin Hakimjavadi, Sophia George).

## Abstract

**Background:** The vaginal microbiome is an underexplored environment which may yield insights about endometrial cancer (EC) pathophysiology and serve as an early marker of disease. Our objective was to evaluate the preoperative vaginal microbiome in women undergoing hysterectomy for benign conditions and EC to identify patterns which could segregate benign disease from EC, and to assess if the microbiome distinguishes low-grade (LG) from high-grade (HG) histologies.

**Methods:** Vaginal microbiome samples were prospectively collected at the time of hysterectomy, and clinicopathologic data collected. Extracted DNA underwent shotgun metagenomics sequencing. Microbial diversity was calculated using the Shannon Index (α-diversity) and PERMANOVA (β-diversity). Hierarchical clustering was used to describe community state types (CST), which were then compared by microbial diversity and grade. Machine learning was utilized to assess the predictive value of grade and histology based on relative bacterial abundance.

**Results:** 61 patients participated: 1) benign gynecologic disease (n=11), 2) LG EC (n=30), and 3) HG EC (n=20). 40 (62.5%) were White, 22 (34.3%) were Black, and 37 (57.8%) were Hispanic. Both α- and β-diversity were associated with tumor grade (p□=□0.026 and p□=□0.035, respectively). Four CST were identified that associated with grade of disease (p=0.036). Different histologies demonstrated variation in CST even within tumor grades (p=0.017). Markers at the species level informed models that predicted benign vs cancer (AUC 0.878), HG versus benign (AUC=0.803), and HG versus LG (AUC=0.771) with high accuracy.

**Conclusions:** The vaginal microbiome segregates benign disease from EC, and is predictive of histology and grade, suggesting it may be an effective tool for screening.

## Introduction

Endometrial cancer is the most common gynecologic malignancy in the United States (1). The incidence of this disease has been increasing, and it is now listed as one of the leading causes of cancer death in women (1, 2). Aside from stratification by epidemiologic risk factors and genetic predisposition (3), there are no routine screening practices for endometrial cancer and women usually present when they develop symptoms. Unfortunately, knowledge of these symptoms is generally poor (4), potentially putting patients at risk for prolonged periods before oncologic evaluation.

While genomic classifications have been suggested to better distinguish between subtypes of endometrial malignancies (5), due to restrictions in cost and expediency required for treatment initiation, the histologic characterization of disease finds more clinical relevance in practice. Type I, or low-grade tumors, are driven by an overabundance of estrogen. These primarily glandular tumors with endometrioid histologies are commonly symptomatic early, diagnosed at earlier stages, and can in many cases be successfully treated with surgery alone. In contrast, type II, or high-grade tumors are characterized by aggressive presentations, often with metastatic disease at diagnosis as they may not be symptomatic in early stages (6). Comprised predominantly of serous, clear cell, carcinosarcoma, high-grade endometrioid, and undifferentiated histologies, type II malignancies have worse survival.

The human microbiome has been shown to influence cancer risk and outcomes by a number of mechanisms, including influencing inflammation, altering the genomic stability in host cells, and producing oncometabolites (7). Defining the microbiome by community state types (CST), which are groups of microbes of similar phyla and abundance, has been useful to describe differences across groups of women, but utilization of CSTs to assess clinical endpoints in endometrial cancer has not previously been described. Because endometrial cancer is a heterogeneous disease comprised of differing histologies and biologic drivers of malignant transformation, comparisons of microbial communities relative to specific histologies and grades may vary and suggest additional unexplored pathways for disease pathogenesis and propagation. Our primary objective was to conduct an exploratory analysis to characterize the preoperative vaginal microbiome in women undergoing surgery for endometrial cancer using metagenomic analyses. The secondary objective was to identify patterns which would reliably segregate not just benign from malignant disease, but also distinguish low-grade from high-grade tumors, as guided by CSTs. Such data may identify opportunities for utilization of the vaginal microbiome in the screening of endometrial cancer, and suggest alternative, unexplored mechanisms of disease pathogenesis.

## Materials and Methods

### Ethical Approval and Consent

Approval for this study was provided by the Institutional Review Board at the University of Miami (protocol #20170660). Informed consent was obtained from all participating patients, with forms provided in English, Spanish, and Haitian Creole. This cross-sectional study is reported in accordance with the Strengthening the Reporting of Genetic Association Studies (STREGA) reporting guideline (8). Patients were consented between February 2018 and October 2018 in a sequential manner without any preplanned stratification. The initial protocol called for an over-sampling of uterine serous carcinoma (planned accrual n=10).

### Population for Study and Patient-related information

Three groups of patients were recruited for the study: 1) Women with benign gynecologic disease undergoing elective surgery for non-malignant conditions, such as fibroids or endometriosis; 2) Women with low-grade (LG) endometrial carcinoma (EC), defined as endometrial intraepithelial neoplasia (EIN), grade 1 or grade 2 endometrioid adenocarcinoma on preoperative endometrial biopsy or uterine curettage; 3) Women with high-grade (HG) endometrial carcinoma, defined as grade 3 endometrioid, serous, small cell, clear cell, undifferentiated, or dedifferentiated carcinoma, or uterine carcinosarcoma, on preoperative endometrial biopsy or uterine curettage. Patients were enrolled sequentially with no planned matching.

Women were required to be ≥18 years of age, able to provide written consent, and able to read and understand English, Spanish, or Haitian Creole. All patients underwent surgery at one of the hospitals affiliated with the physician practice: University of Miami Hospital (UMH), Sylvester Comprehensive Cancer Center (SCCC), or Jackson Memorial Hospital (JMH). Patients were excluded if they had an active gynecologic infection on physician assessment, had any contraindication to the introduction of a swab into the vagina (e.g. severe vaginal stenosis), administration of neoadjuvant chemotherapy, douching within 14 days of surgery, use of vaginal cream or lubricant within 14 days of surgery, or sexual intercourse within five days of surgery.

Patient-specific information was collected from the electronic medical record, including: age at diagnosis, race, ethnicity, final histologic diagnosis, tobacco use, body mass index (BMI), presence of lymphovascular space invasion (LVSI), and results of high-risk HPV DNA testing from most recent Pap smear.

### Specimen collection and processing

On the day of surgery, following induction of anesthesia, and prior to both vaginal preparation with betadine/chlorhexidine and administration of prophylactic antibiotics, the vaginal swab (4N6FLOQSwab, ThermoFisher, #4473979) was placed into the vagina by the attending physician, with care to ensure contact of the swab with the cervix, posterior fornix of the vagina, and vaginal sidewalls. The swab was immediately transferred to the bead tubes which were then snap frozen and kept at –80C. Microbial DNA was extracted with the PureLink microbiome DNA purification kit (ThermoFisher, Invitrogen, #A29790) following manufacturer’s protocol. DNA was eluted in 50ul of AE buffer and quantified using a NanoDrop 2000c Spectrophotometer (Thermo Fisher). Additional details regarding DNA library construction and sequencing can be found in **S1**.

### Statistical analyses

Statistical analyses were performed using custom scripts written in the statistical language R for Statistical Computing. To avoid bias, all patients were included in the analyses, even when missing specific data points, and all available data were included. Summary statistics were used to describe the entire cohort. Significant differences among patient clinical characteristics were determined using Kruskal-Wallis and Wilcoxon signed rank test. The false discovery rates (FDR) were controlled by applying the Benjamini-Hochberg procedure. All tests were two-sided, with significance set at p<0.05. Explanation of the power calculation can be found in **S1**.

### Alpha and beta microbial diversity

Alpha (α) and Beta (β) diversity are standard ecological measures of microbial diversity representing, respectively, the number of unique taxa per sample and similarity in composition between samples. We calculated the observed number of operational taxonomic units (OTUs) as the α diversity measure for each sample within the tumor type groups after rarefaction. We also calculated the Shannon index as our main α-diversity metric, which was generally concordant with observed number of species. We then fitted a linear model for independent samples. The t-type was used to determine statistical significance. For β diversity, we rarefied the data prior to calculating the various distance measures. To test the association between the covariates and β-diversity measures, we used PERMANOVA, a distance-based analysis of variance method based on permutation. An omnibus test, which is a permutation test taking the minimum of the P-values of individual β-diversity measures as the test statistic, was used to combine multiple sources of association evidence provided by different β-diversity measures and an overall association P-value was reported. Ordination plots were generated using classic multi-dimensional scaling (MDS). Analyses of the effects of covariates is provided in **S2**.

### Vaginal community state typing

Briefly, a matrix of sample dissimilarity was created based on the relative abundance of microbial species in each sample using Bray-Curtis distance method. Community State Types (CST) were generated to classify the vaginal microbial communities, to explore community structure, and to reduce dimensionality based on previous reports (9, 10). Samples were clustered into four CSTs using the dissimilarity matrix as the input and Ward’s hierarchical clustering as the method, which minimized the total within-cluster variance. We used gap statistics to determine the optimum number of clusters in the dataset. Considering the sample size, we used k=4 as the optimum number of clusters.

### Differential abundance analysis

We performed microbiome□wide analysis to identify phylum, family, genus, and species that were differentially abundant between samples with different tumor grades and histology. Using phyloseq_to_deseq2 from phyloseq package (11) we transformed microbial relative abundance data into a DESeq dataset with dispersions estimated. We then identified differentially abundant taxa species using the Wald tests from R package DESeq2. We used samples’ species abundance without rarefying to account for variability in read depth between samples. Reported *p*-values were adjusted by the false discovery rate (*p*-adj□<□0.05)

### Gene Expression and Pathway Analysis

We used VIRGO (9) to identify and quantify community gene content, or gene richness, defined as the abundance of non-redundant genes. Non-redundant genes were also annotated with a rich set of functional descriptions. For gene set enrichment analysis (GSEA)(12) we conducted enrichment analysis after constructing gene sets: over-representation and under-representation analyses across pathologies: benign, low-grade EC, high-grade EC and tumor vs benign. We ranked genes based on their fold change between two sample groups using DEseq2 (13). Then using the fgsea R□package, we performed GSEA with three gene sets including KEGG (14), Gene Ontology (GO) (15), and EggNOG, (v.5) (16). Significantly enriched gene sets were filtered based on a cutoff of *q*□<□0.01.

### Machine Learning for Biomarker Discovery

Construction and evaluation of machine learning models on the basis of microbial species was performed using SIAMCAT (17). Read counts at the species level were converted to relative abundances. Species with an overall abundance lower than 0.01 were removed. The statistical association between species and tumor types (or historical types) was measured using Wilcoxon test. FDR was used to correct for multiple testing. Generalized fold change and the prevalence shift between samples with different tumor (or histological) types were calculated for each species.

## Results

### Demographics

The clinical and demographic characteristics of the studied cohort are displayed in **Table 1**. Patients with HG-EC were older than LG-EC and benign patients (q=0.024). There was a significant difference in BMI between benign, HG-EC, and LG-EC patients (q=0.041). More non-Hispanic patients were in the HG-EC cohort versus the LG-EC, in which there were more women of Hispanic ethnicity (q=0.036). There were no differences in tobacco use, human papillomavirus (HPV) status, or race across the three groups (all p>0.05).

**Table 1.** Clinical and demographic characteristics of the cohort^1^.

|  | **Benign**<br>**(n=11)** | **Low-grade**<br>**EC (n=30)** | **High-grade**<br>**EC (n=20)** | **q-value** |
| --- | --- | --- | --- | --- |
|  | Number (%) | Number (%) | Number (%) |  |
| **Age at Surgery** | 51.54 ±10.77 | 60.00 ±11.51 | 61.89 ±10.46 | 0.024 |
| **Body Mass Index** | 30.99 ± 5.46 | 32.77 ± 7.09 | 37.30 ± 7.72 | 0.041 |
| **Tobacco Use** |  |  |  | 0.652 |
| **Current** | 2 (3.4%) | 4 (6.9%) | 3 (5.2%) |  |
| **Former** | 2 (3.4%) | 6 (10.3%) | 1 (1.7%) |  |
| **Never** | 7 (12.1%) | 18 (31%) | 15 (25.9%) |  |
| **Human**<br>**Papillomavirus**<br>**status** |  |  |  | 0.446 |
| **Negative** | 2 (3.4%) | 15 (25.9%) | 6 (10.3%) |  |
| **Positive** | 1 (1.7%) | 1 (1.7%) | 1 (1.7%) |  |
| **Unknown** | 8 (13.8%) | 12 (20.7%) | 12 (20.7%) |  |
| **Ethnicity** |  |  |  | 0.036 |
| **Hispanic** | 6 (9.8%) | 17 (27.9%) | 3 (4.9%) |  |
| **Non-Hispanic** | 5 (8.2%) | 13 (21.3%) | 17 (27.9%) |  |
| **Race** |  |  |  | 0.446 |
| **Asian** | 0 | 0 | 1 (1.7%) |  |
| **Black** | 2 (3.3%) | 10 (16.7%) | 10 (16.7%) |  |
| **White** | 9 (15%) | 19 (31.7%) | 9 (15%) |  |

<sup>1</sup> Parenthetical percentage are relative to entire study cohort. Due to missing data, percentages may not add up to 100%. EC=Endometrial Cancer

### Composition of the vaginal microbiome

The metagenomics data resulted in 4.8% of sequence reads as non-human, of which 64% were taxonomically assigned to 4 major groups: bacteria, archaea, fungi, and viruses. The samples contained 237 bacterial species, representing a majority of previously described vaginal species(18). The most abundant phyla in all samples were *Firmicutes, Actinobacteria, and Bacteroidetes* (**Fig 1**). The most abundant species (based on total abundance over all samples) were *Gardnerella vaginalis, Lactobacillus iners, Streptococcus agalactiae*, and *Lactobacillus gasseri*. The most prevalent (proportion) species (present in all samples) were *Candidatus pelagibacter, Fusobacterium ulcerans, Gardnerella vaginalis*, and *Lactobacillus gasseri*. There was a significantly greater abundance of *Fusobacterium nucleatum* in HG relative to benign samples (log fold-change (FC) 4.3, p=0.02); an increase in the abundance of *Fusobacterium nucleatum* was also seen in LG samples relative to benign, but was not significant (log FC 2.4, p=0.066, **S3**).

**Figure 1.**
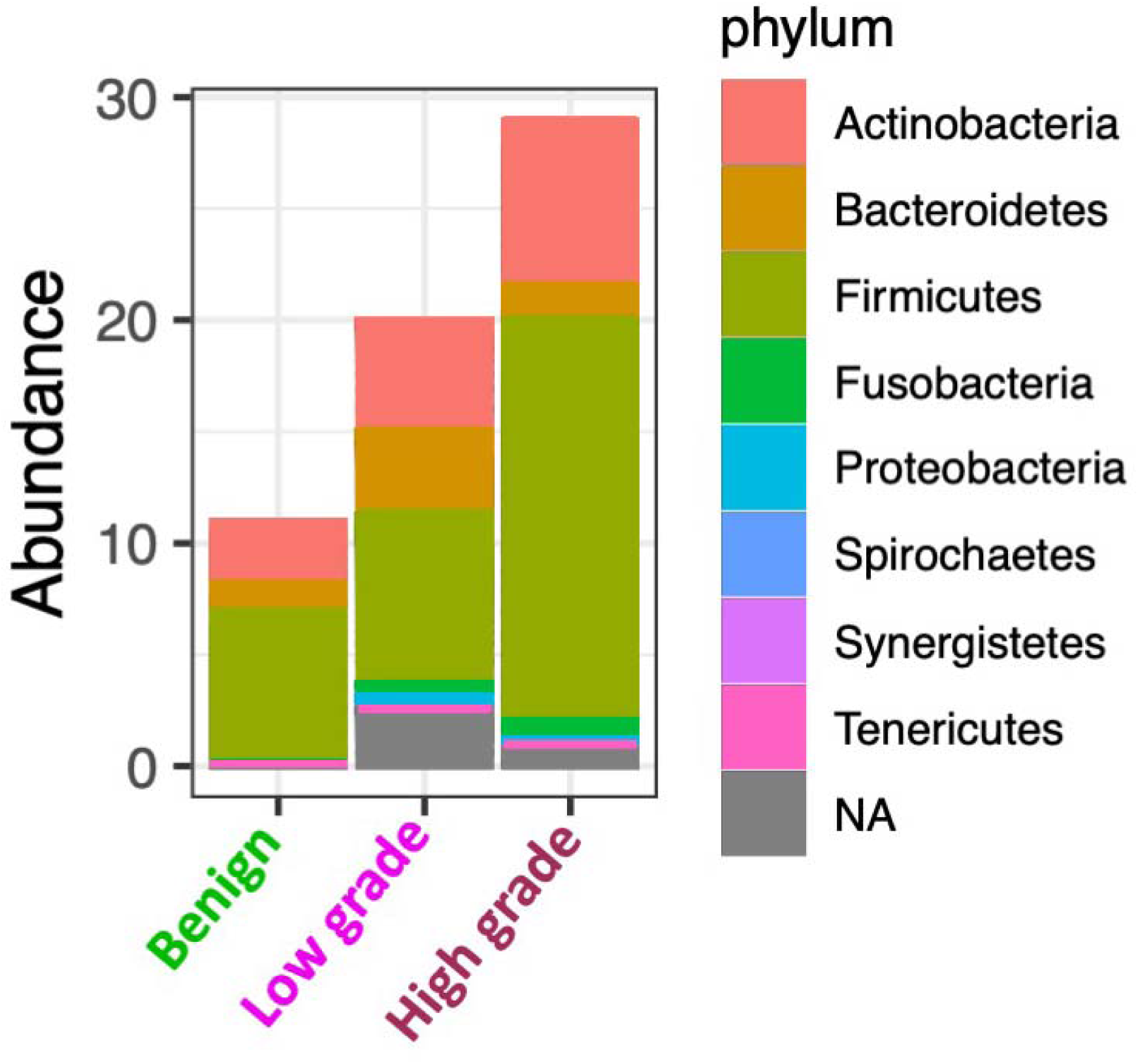
Differential phyla abundance across benign, low-grade EC, and high-grade EC (p=0.093, across all three groups).

### Vaginal microbiota diversity

The vaginal microbiome was significantly different across α- (within samples) and β- (between samples) diversity in patients with the three disease conditions under investigation. Microbial diversity was measured using the Shannon index which accounts for both richness and evenness. Both α- and β-diversity were associated with patient tumor grade (p□=□0.026 and p□=□0.035, respectively).

The α-diversity of LG-EC patients was not significantly different from benign or HG patient samples, however there was a significant increase in diversity from benign to HG disease (q=0.016). This trend suggests that HG disease coincides with a more diverse community of patient’s vaginal microbiome.

The PERMANOVA test was used to quantify beta-diversities based on various clinical variables. Specifically, we evaluated whether tumor types and clinical factors - histology, race, ethnicity, age, BMI and presence of lymphatic vascular invasion (LVSI) - were a significant source of β-diversity, which quantifies dissimilarities of microbial communities based on their composition. Samples grouped by clinical/demographic variables resulted in only one significant difference in microbial diversity (age p=0.049). Meanwhile, both tumor related variables (grade, histology) resulted in significant p-values (grade: p=0.022, histology: p=0.023). This suggests that sample groups are more distinct in their microbial communities based on tumor-related factors than clinical/demographic factors (**S4)**.

### Community State Type Composition and Structure

Four major community state types (CSTs) were identified with significant differences in microbiome composition, diversity, and structure. Each of the four identified CSTs was comprised of communities disproportionately composed by different phyla (**Fig 2A**). *Bacteroidetes* was absent in CST2, and *Fusobacteria* absent in CST1. *Acinetobacteria* and *Firmicules* were variably present across all four CSTs. The most diverse and taxonomically rich cluster was CST4; the least was CST2.

**Figure 2.**
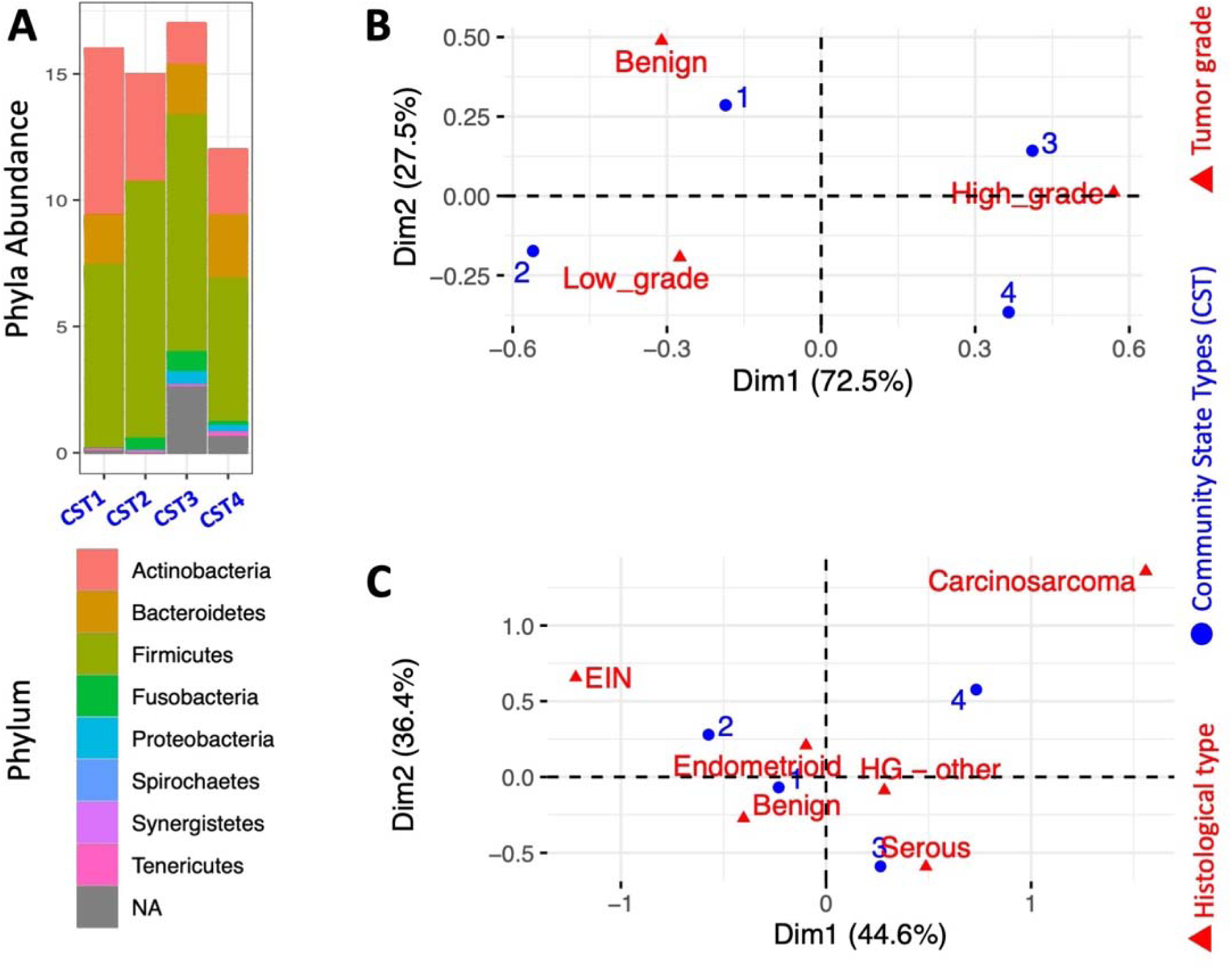
Differential abundance and CST structure. Differences were seen in the microbial phyla abundance by community state type (CST) (A). CSTs were also significantly associated with both tumor grade (B, p=0.036) and tumor histology (C, p=0.017) which ranged from EIN (endometrial intraepithelial neoplasia within the benign category) to endometrioid and rare HG-EC types like serous and carcinosarcoma.

There was statistically significant clustering into CSTs by both grade and histology (**Fig 2B-2C**). Benign disease predominantly clustered in CST1, while LG clustered in CST2, and HG into both CST3 and CST4 (p=0.036). There was also variation in CST clustering by histology (p=0.017). Clinical characteristics and CSTs were evaluated against microbial diversity; only grade and histology had significant associations (**Table 2**).

**Table 2:** Microbial diversities of samples based on clinical variables.

| **Variable** | **Comparison** | **p-value** |
| --- | --- | --- |
| **Age** | <50 years (ref) |  |
|  | 50-69 | 0.13 |
|  | >=70 | 0.39 |
| **Race** | White (ref) |  |
|  | Black | 0.36 |
|  | Asian | 0.61 |
|  | Other | 0.61 |
| **Ethnicity** | Non-Hispanic (ref) |  |
|  | Hispanic | 0.058 |
| **BMI** | <25 (ref) |  |
|  | 25-<30 | 0.69 |
|  | 30-<35 | 0.94 |
|  | 35-<40 | 0.91 |
|  | >=40 | 0.96 |
| **Tumor grade** | Benign (ref) |  |
|  | Low-grade | 0.467 |
|  | High-grade | 0.019 |
| **Histology** | Benign (ref) |  |
|  | EIN | 0.037 |
|  | Endometrioid | 0.229 |
|  | Serous | 0.068 |
|  | Carcinosarcoma | 0.037 |
|  | Other high-grade | 0.116 |
| **LVSI** | Absent (ref) |  |
|  | Present | 0.45 |
| **CST** | 1 (ref) |  |
|  | 2 | 0.00015 |
|  | 3 | 0.12766 |
|  | 4 | 0.00570 |

### Differential abundance analysis

Differential abundance (DA) analyses were conducted to determine the vaginal microbial species enriched or depleted consistently in EC communities. The comparison of average relative abundance between benign versus tumor (LG + HG) revealed that profiles obtained from tumor have only five species with statistically significant DA relative to benign samples (**Fig 3, S5**). Dividing tumor samples into LG and HG profiles identified 30 DA species between HG and LG as well as 17 DA species between HG and benign samples. Noticeably the abundance of 46 species is significantly lower in the HG sample compared to other sample groups. However, two species (*Fusobacterium ulcerans* and *Prevotella bivia*) were found with higher abundance in HG samples. Between LG and benign groups, there were 5 species with significantly greater abundance in the tumors; only *Staphylococcus epidermidis* demonstrated lower abundance.

**Figure 3.**
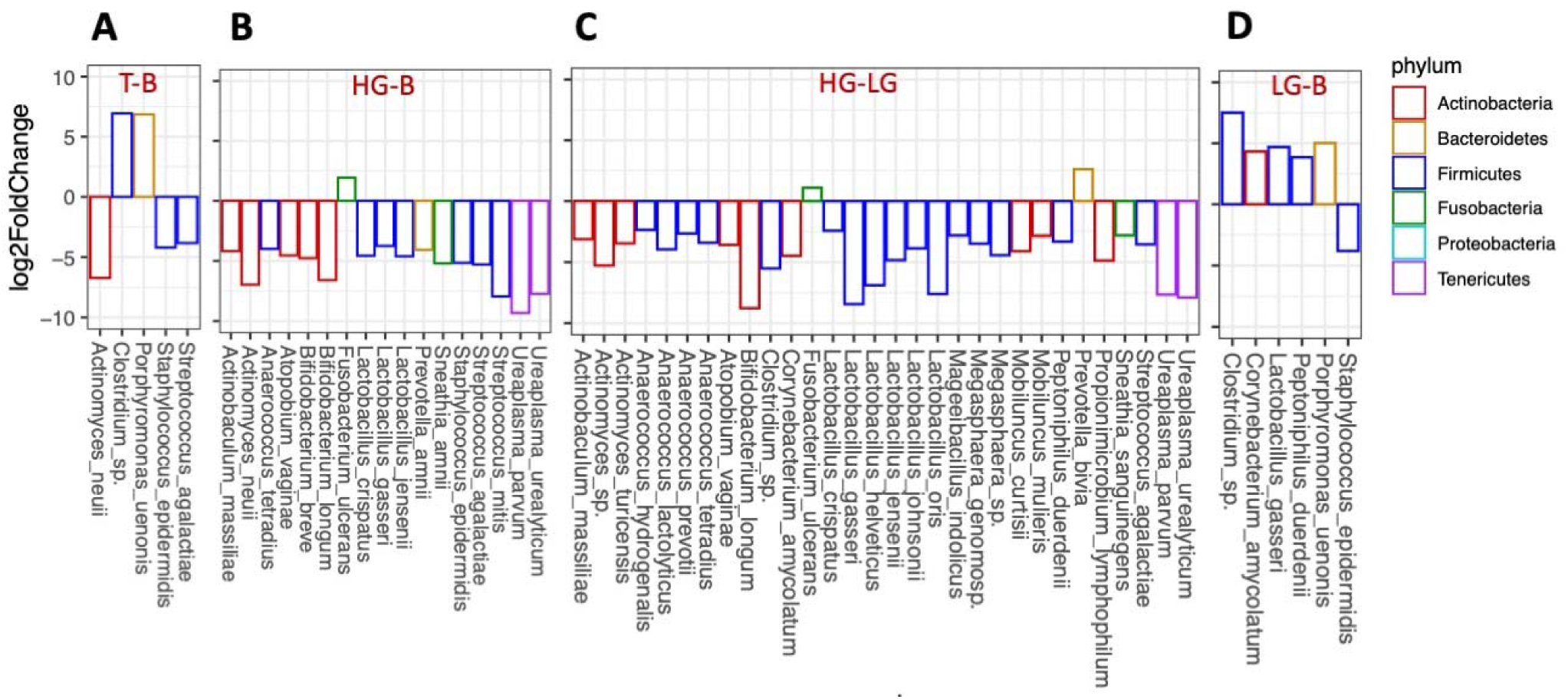
Differential abundance by tumor grade. Positive fold change indicates enrichment of species, whereas negative fold change indicates paucity of species. In both HG and LG tumor vs benign, *Clostridium sp*. and *Porphyromoas uenonis* is more abundant (A). In HG vs benign states *Fusobacterium ulcerans* is the most abundant species (B). Similarly in HG vs LG endometrial cancers, *Fusobacterium ulcerans* and *Prevotella bivia* are the most abundant species (C). LG endometrial cancer metagenomes vs benign have distinct abundance of *Clostridium sp. Corynebacterium amycolatum, Lactobacillus gasseri* and *Peptoniphilus duerdeni*, including *Porphyromoas uenonis* (D). Only taxa with significant changes in abundance are shown (adjusted p<0.05).

### Gene expression and pathway analyses

The metagenomic approach used allows us to investigate gene abundance and thus pathway analyses of the microbiota observed across endometrial pathologies and endometrial cancer histotypes. The HG communities were typically categorized as low gene count as 73.8% of them had less than 1000 genes. Benign communities commonly displayed high gene count as 65% of them had more than 1000 genes. Hierarchical clustering of the profiles was performed using ward linkage based on their Euclidean distance, the result of correspondence analysis conducted for gene richness and diagnosis. We found a strong dependence between the three gene-based clusters and the three tumor grades (p.value =0.025, Chi-square test). The gene-based clusters, however, were independent of other clinical variables including race, ethnicity, BMI groups, age, and disease stage (p.values: 0.64, 0.37, 0.37, 0.08, and 0.22 respectively)(**S6, S7)**.

Using VIRGO, each non-redundant gene was taxonomically and functionally annotated. We next identified significant associations (FDR p < 0.05) between microbial abundance, gene family, and pathway abundance first across tumor and benign, and then more specifically across benign, LG-EC and HG-EC. Kegg pathway analysis of tumor versus benign had the highest number of p<0.05 statically significant associations. Purine metabolism and ABC transporter pathways were up-regulated in tumors whereas genes associated with viral myocarditis, aminoacyl-tRNA biosynthesis and glutathione metabolism were down-regulated in the endometrial tumor microbiota (**S8A**). Additional analyses of the metagenome of HG endometrial cancers alone compared to benign revealed the only pathway significantly upregulated is biosynthesis of siderophore group non-ribosomal peptides. Conversely, pathways downregulated included the pyrimidine metabolism, purine metabolism, homologous recombination, and ABC transporters (**S8B)**. The downregulation of gene sets in homologous recombination, mismatch repair and ABC transporters was unique to HG-EC microbiota while ABC transporter pathway was up in the combination tumors.

### Biomarker Discovery

To examine the diagnostic value of the vaginal microbiome, we constructed random forest (RF) models that could specifically classify samples according to patients’ tumor types including a) benign vs. tumor samples b) HG tumors vs. benign, and c) HG tumors vs LG tumors. To detect useful species markers of tumor, we conducted a fivefold cross-validation on a random forest model between case and control samples in the discovery phase. For each model a different set of species was identified as an optimum microbiome signature, consisting of a various number of features and performance of the constructed models based on the area under the receiver operating characteristic (ROC) curve (**Fig 4A,4C,4E**). The tumor vs benign model selected 3 important species. The discriminant model based on the abundance of these species effectively distinguishes tumor from benign disease (mean prediction AUC = 0.878, **Fig 4B**). Two other RF models generated from additional species abundance distinguished HG from benign, and LG from HG with AUC of 0.80 and 0.77 respectively (**Fig 4D-F**).

**Figure 4.**
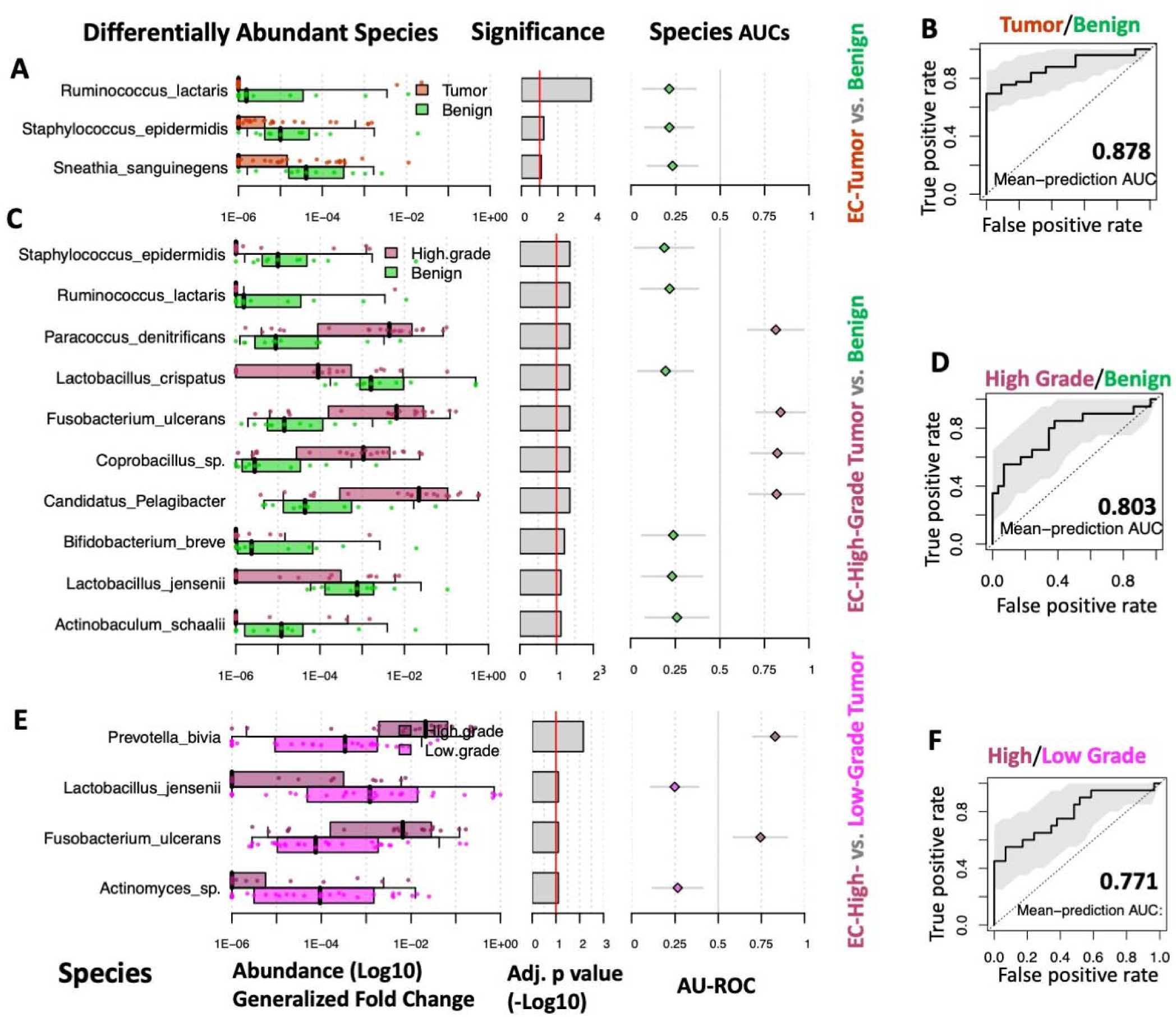
Biomarker Discovery by grade. Validation was performed on random forest classifier models, which identified an optimal microbiome signature for each cohort (A, C, E). These signatures were used to construct receiver operating curves which discriminate benign versus tumor (B), HG versus benign (D), and HG versus LG (F).

We examined the performance of models trained by samples labeled according to histologic subtype (e.g. serous, endometrioid) (**S9**). The highest prediction performance obtained from the model that trained to distinguish benign samples from samples labeled as serous EC (mean AUC=0.826) followed by two models that distinguish benign from endometrioid samples (mean AUC=0.795) and serous from endometrioid (mean AUC=0.776). Each of these three histologic classifier models are based 50, 60, and 65 biomarker species, respectively.

## Discussion

Among patients with EC, the vaginal microbiome demonstrates significant variation by tumor pathologic characteristics. This investigation establishes that not only do prominent species vary by grade, but so too do microbial abundance and CST. These findings represent a novel perspective on the microbial content of the vagina and how the confluence with the uterus may provide opportunities for EC screening and further exploration of the role of the microbiome in disease development.

There have been few studies about the vaginal microbiome in patients with EC. In 2016, Walther-Antonio et al assessed the microbiome (16S) of the entire gynecologic tract of 17 patients with EC and 10 with benign uterine conditions. The authors reported that the microbiome across different gynecologic sites was significantly correlated, suggesting that vaginal sampling is an accurate surrogate of the microbiome within the uterus. Subsequent investigations have also confirmed that the vaginal microbiome mirrors that of the upper genital tract among women with cancer (19). Additionally, it was reported that the pattern of presence/absence of *Porphyromonas* and *Atopobium* species was predictive of EC (AUC 0.90)(20). Within our cohort, neither of these phyla demonstrated significant abundance. In contrast to the Walther-Antonio study, however, we assessed specific tumor grade relative to benign conditions. Additionally in their study all EC patients were White; 37% of our population was Black. Microbial diversity has been shown to be greater in Black versus White women (21), and CST in premenopausal women defined by Lactobacillus varies across all races (Black versus White versus Asian)(22), so the differences in the populations between our two studies may account for the discrepancy.

A follow-up study by Walsh et al. which included 56 patients with endometrioid histology and 10 with non-endometrioid, high-grade, histology, also reported that *Porphyromonas somerae* was a predictive biomarker in EC, and that additional sensitivity to disease detection was added by including patient-specific factors such as BMI, vaginal pH, and menopausal status (19). While we did not assess vaginal pH in the current study, we found no association between age or BMI and microbial diversity (Table 2). Our methodology, however, differed in that our data were segregated categorically to represent clinically meaningful groups (i.e. BMI following WHO categorization; age of 50 serving as a surrogate for menopause). This variation in analysis may account for our findings, but may again be reflective of differences in the population of study relative to our own, as 97% of the Walsh cohort was White and only 10 patients had HG cancers. As microbial diversity in the current study was associated with tumor factors only (grade and histology) and not with categorical clinical factors, it suggests that patient-specific factors may not necessarily need to be included in a predictive model for screening.

While the differential phyla abundance between benign and tumor provides some insight into the local vaginal environment, differences in species abundance may also be meaningful in terms of tumor pathogenesis. *Prevotella bivia*, with greater than a 6-fold abundance in HG vs LG, is associated with pelvic inflammatory disease and bacterial vaginosis. *P. bivia* has been shown to upregulate proinflammatory (LAMP3, STAT1 and TAP1) genes in cervical cancer (23). Furthermore, Lactobacillus spp, which are underrepresented in HG vs benign and HG vs LG, are known to inhibit *P. bivia* (24). *Bifidobacterium longum* was the most greatly suppressed species in terms of abundance in HG vs LG disease. *B. longum* has been shown to have low relative abundance in patients with the most aggressive forms of gastric cancer, suggesting it may be protective (25). It has also been shown to improve immune-mediated tumor control (26). *Fusobacterium ulcerans* also demonstrated higher abundance in HG. This species has an association with cellular ulceration by secretion of high levels of butyrate (27); very little data exist about its role in cancer pathogenesis. *Fusobacterium nucleatum*, though not one of the most abundant species contributing to the predictive models, but with a greater than 4-fold presence in HG vs benign, has been found to promote tumor growth (28), associate with high microsatellite instability (29), and induce chemotherapy resistance (30). Patients with cervical cancer who have high levels of intratumoral *F. nucleatum* have worse progression free and overall survival (31). In colorectal cancer (CRC), the bacterium secretes the adhesin Fap2, which binds to galactose *N-*acetyl-D-galactosamine (Gal-GalNAc), facilitating the enrichment of tumor cells (30). Gal-GalNAc levels have been shown to be higher in uterine adenocarcinomas relative to benign endometrium (32), and over-expression of the transferases that facilitate Gal-GalNAc glycosylation are strongly associated with histologic grade of tumor and myometrial invasion (33). In CRC cells in vitro, a high abundance of intratumoral *F. nucleatum* also activates autophagy, thus inducing resistance to platinum-based chemotherapy (30). The role of all these bacteria in the pathogenesis and treatment of EC, and specifically high-grade histologies, requires further investigation.

The mechanisms by which the microbiome influences EC pathogenesis have yet to be determined but are likely multifactorial in the context of tumor stromal function and alterations in cancer cell signaling pathways. Lu et al recently reported that the presence of specific bacteria in the endometrium are associated with variable levels of the pro-inflammatory cytokines IL-6, IL-8, and IL-17 (34). These molecules are known to modify the local microenvironment, and have been implicated in gynecologic cancer development through increased angiogenesis, cellular proliferation, and modification of local immune response (35-37). In patients with CRC, the presence of *F. nucleatum*, may activate the Wnt/β-catenin signaling pathway (38). In the endometrium, this pathway is important for normal physiologic cellular proliferation during the menstrual cycle, but oncogenic activation is also associated with EC development (39, 40). Consideration should also be given to environmental mediators of microbial content, as practices such as douching have also been shown to favorably modify the gynecologic tract for pathogens (41).

There are several limitations to our study. Our population was from a single institution, so the results may not be applicable in other study environments. Nonetheless, the population was racially and ethnically diverse, which may increase generalizability. Though our sample population was small, we were still able to identify statistically significant associations between CSTs and histology, with >90% power (**Supplement 1**). Additionally, these relationships were maintained across our analyses, including composition and differential abundance. We designed the study to specifically include more serous carcinoma patients, and this oversampling approach allowed for greater representation of understudied, high-risk endometrial histologies, relative to other reports (19, 20). Moreover these analyses used a metagenomics approach instead of 16S rRNA sequencing in the assessment of EC-associated microbiomes. This allowed for a more robust evaluation of relative microbial abundance and diversity. While others have advocated for the use of one or two species to discriminate between benign and malignant (19, 20), this study included multiple bacterial species to define clusters of organisms that collectively predicted not just malignancy, but subsets of disease. Such an approach can increase the accuracy of these models, especially as consideration is given to the clinical utility of the vaginal microbiome in disease assessment.

## Conclusions

The vaginal microbiome reasonably segregates not only EC from benign disease, but also has strong potential predictive value by grade and histology. Further study in larger populations is needed to define its role as a biomarker in screening, early detection, and discernment between histologies as such data may directly drive changes in clinical decision-making. It will also be important to further characterize the relationships between the microbiome and tumor microenvironment, be they symbiotic or simply associative, and how these may contribute to disease etiology, tumor propagation, and potential novel therapeutic approaches.

## Data Availability

All data associated with this study are available upon reasonable request to the authors and have been uploaded to Gene Expression Omnibus. SRA Submission ID: SUB9784683

## Acknowledgments

The research was supported by the Jay Weiss Institute for Health Disparities Research, the Ruth Helen O’Bryan Wright/Gyn Precision Medicine Initiative, Sylvester Comprehensive Cancer Center, University of Florida Health Cancer Center, National Cancer Institute of the National Institutes of Health under (P30CA240139). We thank Sylvia Daunert and Gregory O’Connor for critical review of the manuscript. The content is solely the responsibility of the authors and does not necessarily represent the official views of the National Institutes of Health.

## Data and materials availability

All data associated with this study are available upon request and have been uploaded to Gene Expression Omnibus. SRA Submission ID: SUB9784683

Title: Vaginal microbiome predicts endometrial cancer grade and histology

## Author Contributions

Hesamedin Hakimjavadi – methodology, software, validation, formal analysis, data curation, writing-original draft, writing-review&editing

Sophia George – conceptualization, methodology, investigation, resources, writing-original draft, writing-review&editing, supervision, project administration

Michael Taub – investigation, writing-review&editing

Leah Dodds – investigation, writing-review&editing

Alex Sanchez-Covarrubias - investigation, writing-review&editing

Marilyn Huang, MD - investigation, writing-review&editing

Matthew Pearson - investigation, writing-review&editing

Brian Slomovitz - investigation, writing-review&editing

Erin Kobetz – methodology, writing-review&editing

Raad Z. Gharaibeh – software, formal analysis, writing-review&editing

Ramlogan Sowamber - investigation, writing-review&editing

Andre Pinto - investigation, writing-review&editing

Srikar Chamala - methodology, software, validation, formal analysis, data curation, writing-original draft, writing-review&editing

Matthew Schlumbrecht – conceptualization, methodology, validation, writing-original draft, writing-review&editing, supervision, project administration, funding acquisition

## References

1. Siegel R, Miller K, Fuchs H, Jemal A. Cancer Statistics, 2021. CA Cancer J Clin. 2021;71(1):7–33.

2. Clarke M, Devesa S, Harvey S, Wentzensen N. Hysterectomy-corrected uterine corpus cancer incidence trends and differences in relative survival reveal racial disparities and rising rates of nonendometrioid cancers. J Clin Oncol. 2019.

3. Lu K, Daniels M. Endometrial and ovarian cancer in women with Lynch syndrome: update in screening and prevention. Fam Cancer. 2013;12(2):273–7.

4. Soliman P, Bassett R, Wilson E, Boyd-Rogers S, Schmeler K, Milam M, et al. Limited public knowledge of obesity and endometrial cancer risk. Obstet Gynecol. 2008;112(4):832–42.

5. Kandoth C, Schultz N, Cherniack A, Akbani R, Liu Y, Shen H, et al. Integrated genomic characterization of endometrial carcinoma. Nature. 2013;497(7447):67–73.

6. Felix A, Weissfeld J, Stone R, Bowser R, Chivukula M, Edwards R, et al. Factors associated with type I and type II endometrial cancer. Cancer Causes Control. 2010;21(11):1851–56.

7. Bultman S. Emerging roles of the microbiome in cancer. Carcinogenesis. 2014;35(2):349–55.

8. Little J, Higgins JP, Ioannidis JP, Moher D, Gagnon F, von Elm E, et al. STrengthening the REporting of Genetic Association Studies (STREGA): an extension of the STROBE statement. PLoS Med. 2009;6(2):e22.

9. Ma B, France M, Crabtree J, Holm J, Humphrys M, Brotman R, et al. A comprehenisve non-redundant gene catalog reveals extensive within-community intraspecies diversity in the human vagina. Nat Commun. 2020;11(1):940.

10. De Seta F, Campisciano G, Zanotta N, Ricci G, Comar M. The vaginal community state type microbiome-immune network as a key factor for bacterial vaginosis and aerobic vaginitis. Front Microbiol. 2019;10:2451.

11. McMurdie P, Holmes S. phyloseq: an R package for reproducible interactive analysis and graphics of microbiome census data. PLoS One. 2013;8(4):e61217.

12. Subramanian A, Tamayo P, Mootha VK, Mukherjee S, Ebert BL, Gillette MA, et al. Gene set enrichment analysis: a knowledge-based approach for interpreting genome-wide expression profiles. Proc Natl Acad Sci U S A. 2005;102(43):15545–50.

13. Love M, Huber W, Anders S. Moderated estimation of fold change and dispersion for RNA-seq data with DESeq2. Genome Biol. 2014;15(550).

14. Kanehisa M, Araki M, Goto S, Hattori M, Hirakawa M, Itoh M, et al. KEGG for linking genomes to life and the environment. Nucleic acids research. 2007;36:D480–84.

15. The Gene Ontology (GO) database and informatics resource. Nucleic acids research.32:D258–61.

16. Jensen L, Julien P, Kuhn M, von Mering C, Muller J, Doerks T, et al. eggNOG: automated construction and annotation of orthologous groups of genes. Nucleic acids research. 2007;36:D250–54.

17. Wirbel J, Zych K, Essex M, Karcher N, Kartal E, Salazar G, et al. Microbiome meta-analysis and cross-disease comparison enabled by the SIAMCAT machine learning toolbox. Genome Biol. 2021;22(1):1–27.

18. Ma B, France MT, Crabtree J, Holm JB, Humphrys MS, Brotman RM, et al. A comprehensive non-redundant gene catalog reveals extensive within-community intraspecies diversity in the human vagina. Nat Commun. 2020;11(1):940.

19. Walsh D, Hokenstad A, Chen J, Sung J, Jenkins G, Chia N, et al. Postmenopause as a key factor in the composition of the Endometrial Cancer Microbiome (ECbiome). Sci Rep. 2019;9(1):19213.

20. Walther-Antonio M, Chen J, Multinu F, Hokenstad A, Distad T, Cheek E, et al. Potential contribution of the uterine microbiome in the development of endometrial cancer. Genome Med. 2016;8:122.

21. Clark LH, Keku TO, McCoy NA, Hawkins G, Bae-Jump VL, Brewster WR. Alterations in the uterine microbiome in patients with early endometrial cancer: Variations by ethnicity and obesity. Journal of Clinical Oncology. 2017;35(15_suppl):e17114–e.

22. France M, Ma B, Gajer P, Brown S, Humphrys M, Holm J, et al. VALENCIA: a nearest centroid classification method for vaginal microbial communities based on composition. Microbiome. 2020;8(1):166.

23. Lam KC, Vyshenska D, Hu J, Rodrigues RR, Nilsen A, Zielke RA, et al. Transkingdom network reveals bacterial players associated with cervical cancer gene expression program. PeerJ. 2018;6:e5590–e.

24. Atassi F, Brassart D, Grob P, Graf F, Servin AL. Lactobacillus strains isolated from the vaginal microbiota of healthy women inhibit Prevotella bivia and Gardnerella vaginalis in coculture and cell culture. FEMS Immunol Med Microbiol. 2006;48(3):424–32.

25. Devi T, Devadas K, George M, Gandhimathi A, Chouhan D, Retnakumar R, et al. Low Bifidobacterium abundance in the lower gut microbiota is associated with Helicobacter pylori-related gastric ulcer and gastric cancer. Front Microbiol. 2021;12:631140.

26. Matson V, Fessler J, Bao R, Chongsuwat T, Zha Y, Alegre M, et al. The commensal microbiome is associated with anti-PD-1 efficacy in metastatic melanoma patients. Science. 2018;359:104–8.

27. Adriaans B, Garelick H. Cytotoxicity of Fusobacterium ulcerans. J Med Microbiol. 1989;29:177–80.

28. Bullman S, Pedamallu C, Sicinska E, Clancy T, Zhang X, Cai D, et al. Analysis of Fusobacterium persistence and antibiotic response in colorectal cancer. Science. 2017;358(6369):1443–48.

29. Mima K, Nishihara R, Qian Z, Cao Y, Sukawa Y, Nowak J, et al. Fusobacterium nucleatum in colorectal carcinoma tissue and patient prognosis. Gut. 2015;65(12).

30. Brennan C, Garrett W. Fusobacterium nucleatum - symbiont, opportunist and oncobacterium. Nat Rev Microbiol. 2019;17(3):156–66.

31. Huang S, Chen J, Lian L, Cai H, Zeng H, Zheng M, et al. Intratumoral levels and prognostic significance of Fusobacterium nucleatum in cervical carcinoma. Aging (Albany NY). 2020;12(22):23337–50.

32. Abed J, Maalouf N, Parhi L, Chaushu S, Mandelboim O, Bachrach G. Tumor targeting by Fusobacterium nucleatum: A pilot study and future perspectives. Front Cell Infect Microbiol. 2017;7(295).

33. Ngyuen T, Kurita T, Koi C, Murakami M, Kagami S, Hachisuga T, et al. GalNAc-T6 in the relationship with invasion ability of endometrial carcinomas and prognostic significance. Am J Cancer Res. 2017;7(5):1188–97.

34. Lu W, He F, Lin Z, Liu S, Tang L, Huang Y, et al. Dysbiosis of the endometrial microbiota and its association with inflammatory cytokines in endometrial cancer. Int J Cancer. 2021;147:1708–16.

35. Che Q, Liu B, Wang F, He Y, Lu W, Liao Y, et al. Interleukin 6 promotes endometrial cancer growth through and autocrine feedback loop involving ERK-NFκB signaling pathway. Biochem Biophys Res Commun. 2014;446(1):167–72.

36. Fujimoto J, Aoki I, Khatun S, Toyoki H, Tamaya T. Clinical implications of expression of interleukin-8 related to myometrial invasion with angiogenesis in uterine endometrial cancers. Ann Oncol. 2002;13(3):430–4.

37. Lai T, Wang K, Hou Q, Zhang J, Yuan J, Yuan L, et al. Interleukin 17 induces up-regulation of chemokine and cytokine expression via activation of the nuclear factor κB and extracellular signal-regulated kinase 1/2 pathways in gynecologic cancer cell lines. Int J Gynecol Cancer. 2011;21(9):1533–39.

38. Rubinstein M, Baik J, Lagana S, Han R, Raab W, Sahoo D, et al. Fusobacterium nucleatum promotes colorectal cancer by inducing Wnt/B-catenin modulator Annexin A1. EMBO Rep. 2019;20(4):e47638.

39. Wang Y, van der Zee M, Fodde R, Blok L. Wnt/B-catenin and sex hormone signaling in endometrial homestasis and cancer. Oncotarget. 2010;1(7):674–84.

40. Kiewisz J, Wasniewski T, Kmiec Z. Participation of WNT and B-catenin in physiological and pathological endometrial changes: Association with angiogenesis. Biomed Res Int. 2015;2015:854056.

41. Seay J, Mandigo M, Kish J, Menard J, Marsh S, Kobetz E. Intravaginal practices are associated with greater odds of high-risk HPV infection in Haitian women. Ethn Health. 2017;22(3):257–65.

