## Supplement 1 for "Vaginal microbiome is associated with endometrial cancer grade and histology"

DNA library construction and sequencing

Extracted vaginal microbial DNA - 1ug for each sample underwent quality control. Metagenomics shotgun sequencing was performed by BGI Americas. Briefly, DNA was sheared into smaller fragments (300-400bp) by Covaris S/E210. Then the overhangs resulting from fragmentation were converted into blunt ends by using T4 DNA polymerase, Klenow Fragment and T4 Polynucleotide Kinase. Adapters were ligated to the ends of the DNA fragments. Desired fragments were purified then selectively enriched and amplified by PCR. The index tag could be introduced into the adapter at the PCR stage as appropriate and we did a library quality test. At last, the qualified BS library would be used for sequencing on BGISEQ PE100 platform. The metagenomic raw reads from all samples were, trimmed, and filtered to the host contamination using KneadData pipeline v0.7.3 in paired end mode with Bowtie2 (https://bitbucket.org/biobakery/kneaddata). Quality controlled metagenome reads then processed with VIRGO (human vaginal non-redundant gene catalog) and profiled for microbial species abundances, gene content, and functions.

Power Calculation

To compute statistical power, we used the statistical package “micropower” available in R. This package quantifies group level effects by the adjusted coefficient of determination, omega-squared (w^2^) at different sample sizes using pairwise distances and PERMANOVA. We downloaded abundance tables from the NIH human microbiome project (HMP) to serve as the input to run the analysis. All samples identified as coming from the genitourinary tract were included (n=65). We simulated a set of weighted Jaccard distances matrices from randomly created OTU tables for which within-group distances match the distribution of distances observed in the HMP genitourinary data set. We run the package three times to assess PERMANOVA power with either 10, 15 or 20 subjects per group. At 90% power, we found that 10 subjects per group detect a w^2^ of 0.038; 15 subjects per group detect a w^2^ of 0.019 and 20 subjects per group detect a w^2^ of 0.010. Effect sizes just below the value of those observed in reference tables provided by Kelly et al. are considered powerful enough with the proposed number of subjects. In our analysis, the w^2^ value observed in 15 subjects per group at 90% power was enough to detect a difference.
