## Supplement 2 for "Vaginal microbiome is associated with endometrial cancer grade and histology"

**Supplement 2: Analysis of Confounders**

Some clinical variables, including patients' age, BMI, and ethnicity, can potentially influence microbiome composition beyond the tumor type. Therefore, we examined these three clinical variables as covariates that have the potential to bias detected statistical differences between

high- and low-grade tumor microbiome.

1. Conditional entropy quantifies the unique information contained in the label variable (low- and high-grade tumors) respective to other likely confounding factors (BMI, Ethnicity, and Age; See the highlighted row). Identical label variables and confounding variables which share the exact same information will have a value of zero. The values in the highlighted cells correlates with unique information provided by BMI, ethnicity, and age.

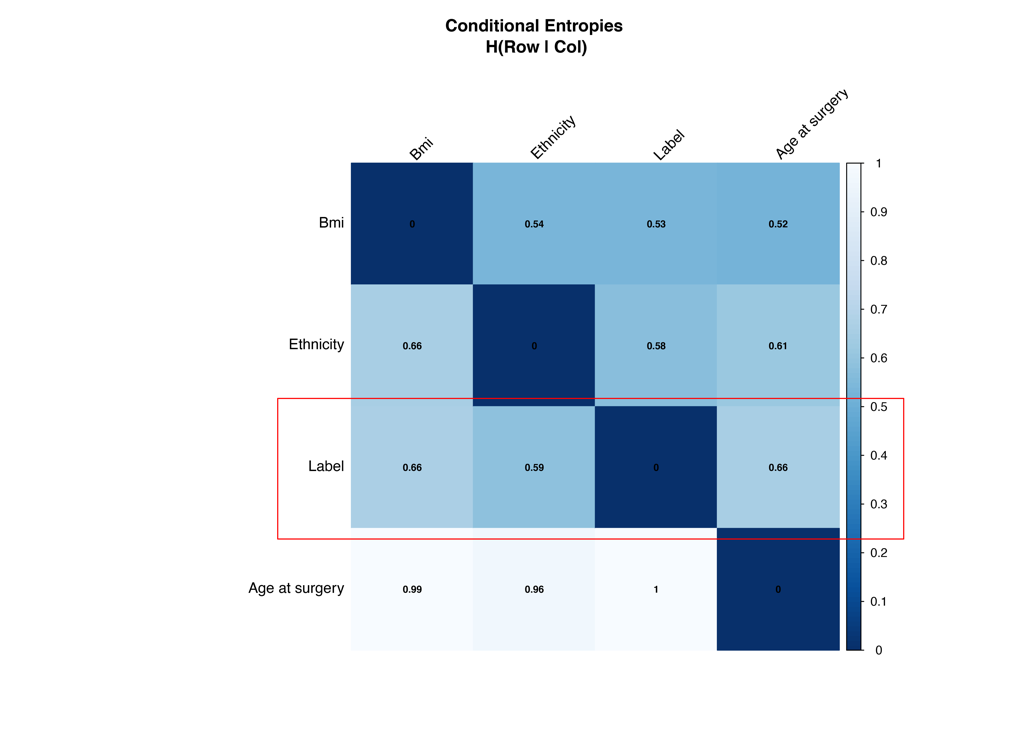

2. Age as covariates does not confound the association between microbiome and tumor type (low- vs. high-grade)

Distribution: The age distribution at surgery is similar between patients with low or high-grade tumors (figs A, B, D). And there is no significant difference between the ages of the two groups (fig. C, E). Analysis of variance (using ranked abundance data) shows more species differ by tumor type than age (dot size is proportional to the mean relative abundance across samples. Therefore, age as a covariate explains a small portion of variations observed in patients' microbiome. This is confirmed by the result of two statistical analyses (Covariate logistic regression and ROC analysis) that indicate tumor samples from the selected group (low-and high-grade pool) cannot be accurately distinguished by age as a covariate (adjusted p.value=0.32; AUROC= 0.61 using SIAMCAT classification workflows).

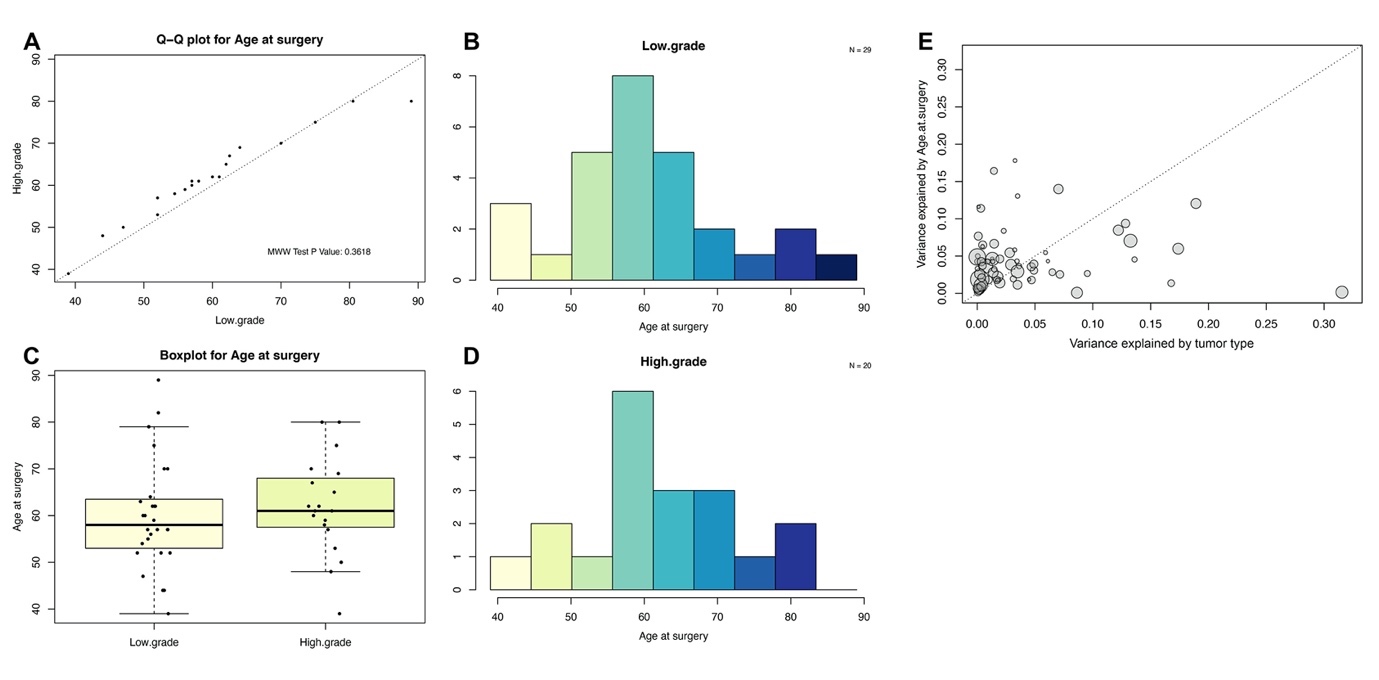

3. Logistic regression and ROC analysis of age, BMI, and ethnicity as covariates that potentially confound the association between microbiome and tumor type. Ethnicity as covariates confound the association between microbiome and tumor type (low- vs. high-grade)

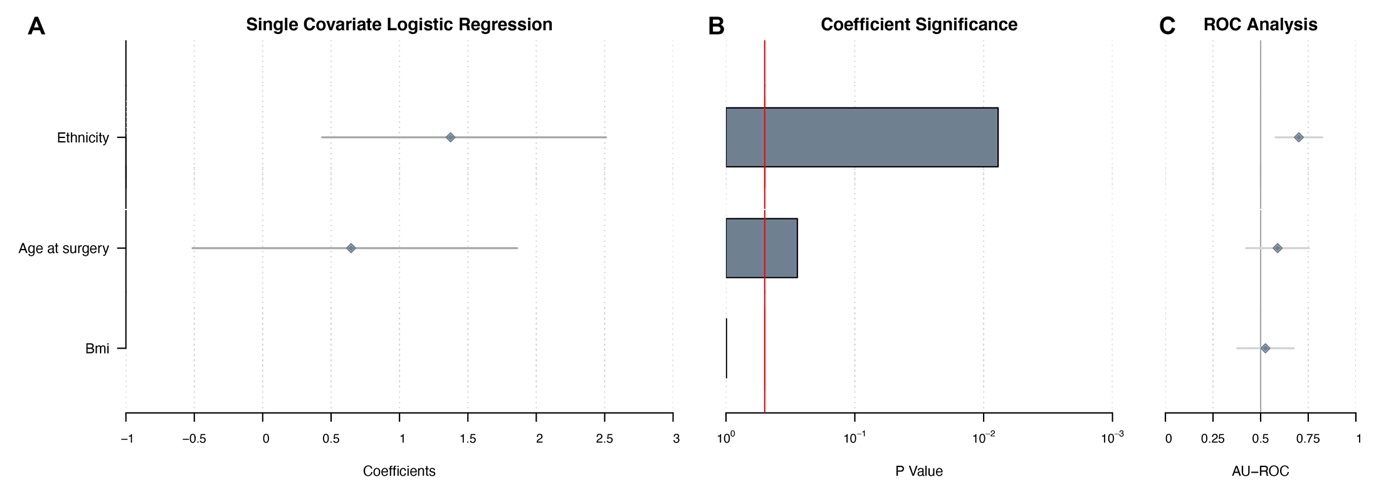

4. BMI as a covariate explains a significant portion of variations observed in patients' microbiome with low- or high-grade tumors but is unlikely to be a confounding factor. Although analysis of variance shows more species differ by BMI than tumor type (Fig. E), the BMI distribution at surgery is similar between patients with low or high-grade tumors (figs A, B, D). Furthermore, there is no significant difference between the ages of the two groups (fig. C). Statistical analyses also indicates that tumor samples from the selected group (low-and high-grade pool) cannot be accurately distinguished by BMI as a covariate (adjusted p.value=1; AUROC= 0.53). Therefore, BMI as covariates is unlikely to confound the association between microbiome and tumor type (in low- and high-grade samples)

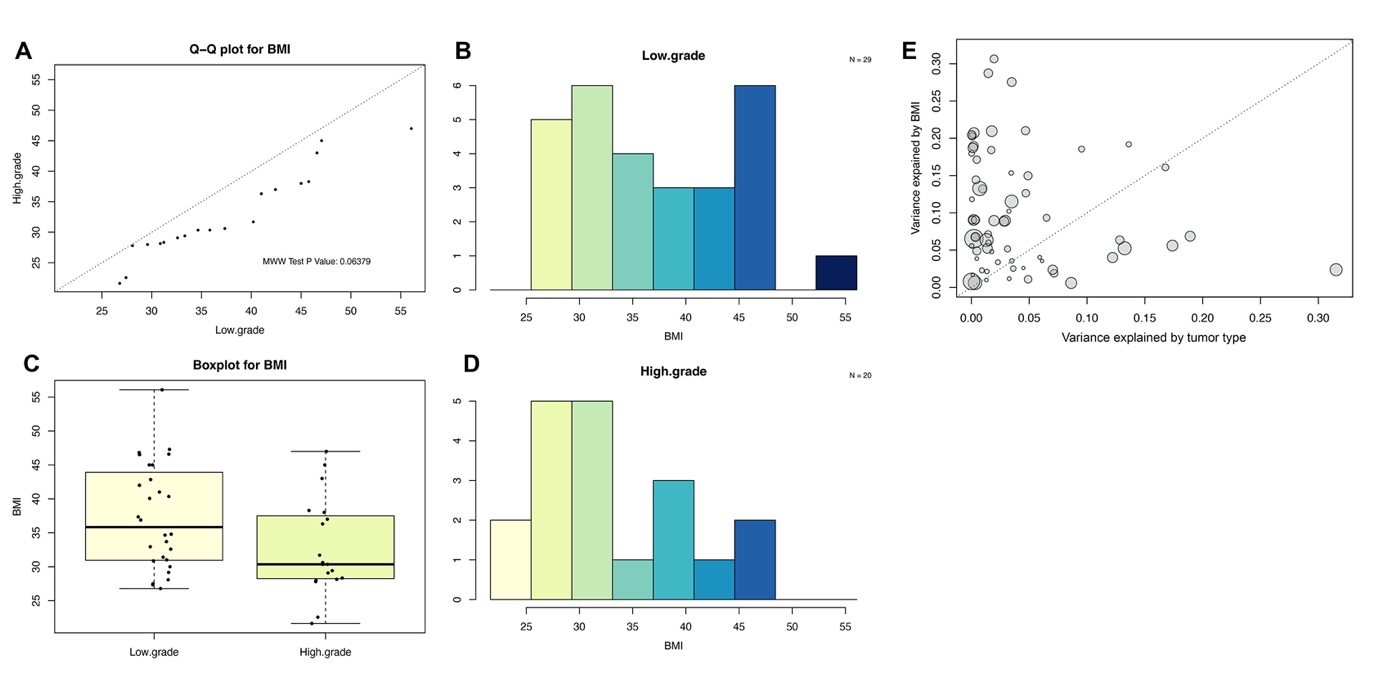

5. Age as covariates confound the association between microbiome and tumor type (low- vs. high-grade)

Distribution: The ethnicity distribution at surgery significantly different between patients with low or high-grade tumors (p.value= 0.006. see figs A,B).

Analysis of variance shows most of species differ by tumor type rather than ethnicity.

Therefore, ethnicity as a covariate explains a small portion of variations observed in patients' microbiome.

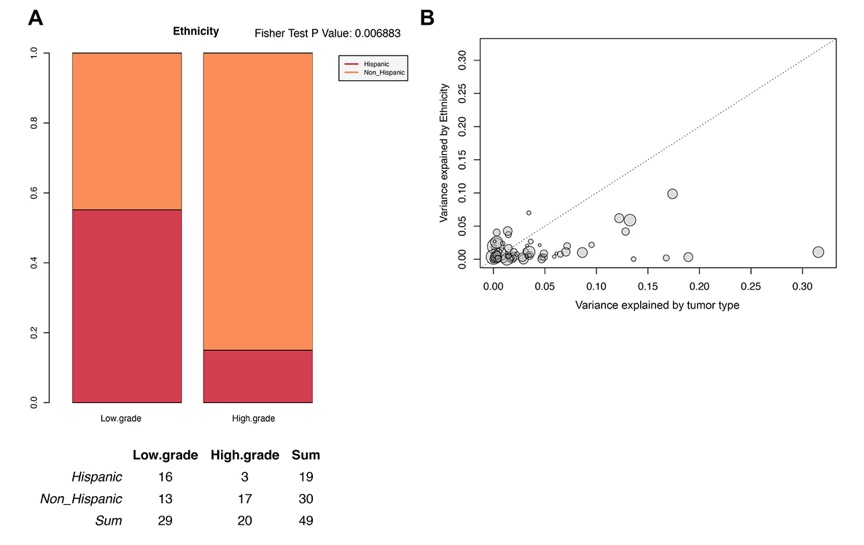
