## Supplementary figures and images for "Vaginal microbiome is associated with endometrial cancer grade and histology"

### Supplement 3

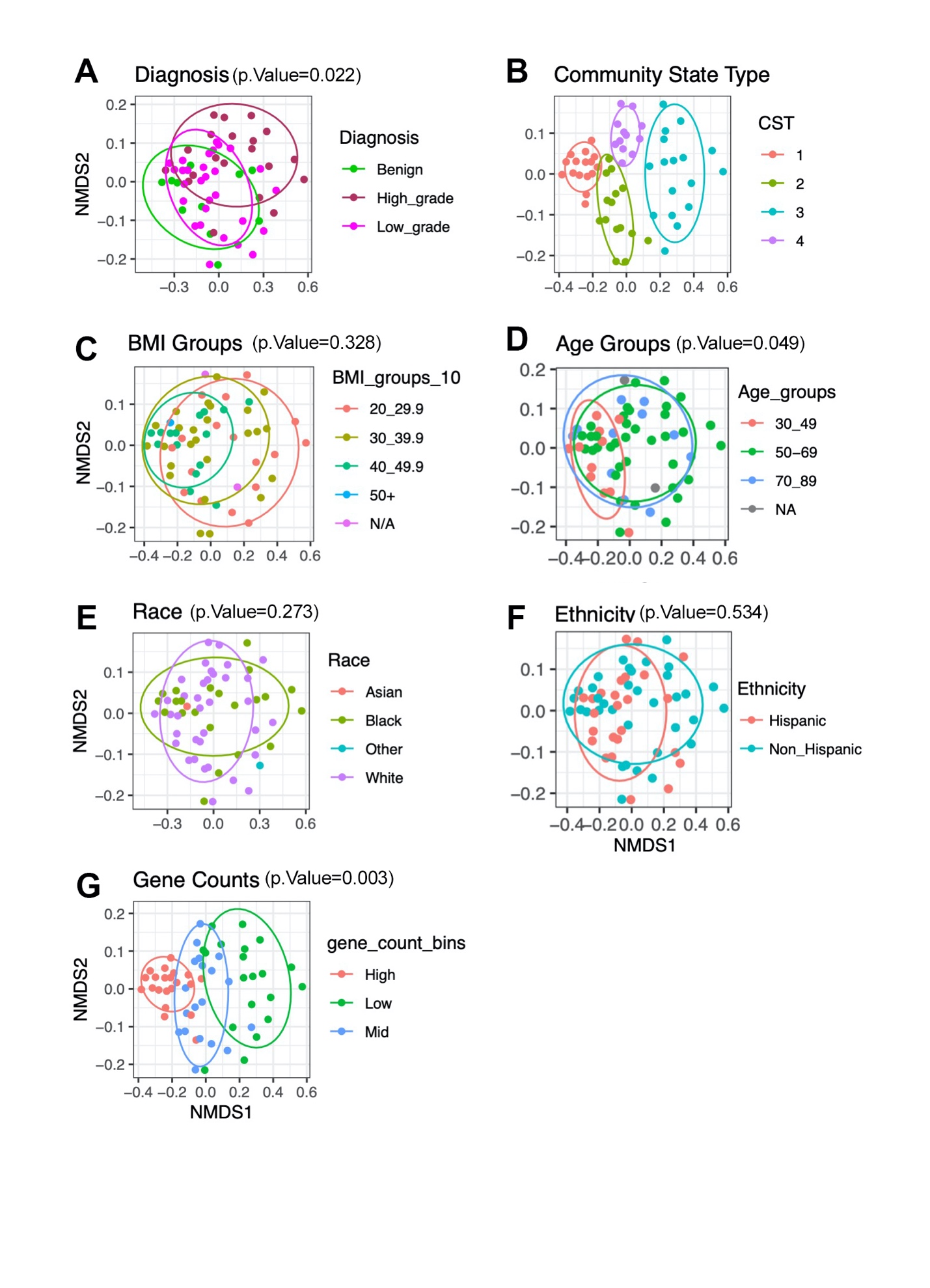

**Supplement 4**. PCA visualization of beta diversity based on seven different clinical variables.

### Supplement 4

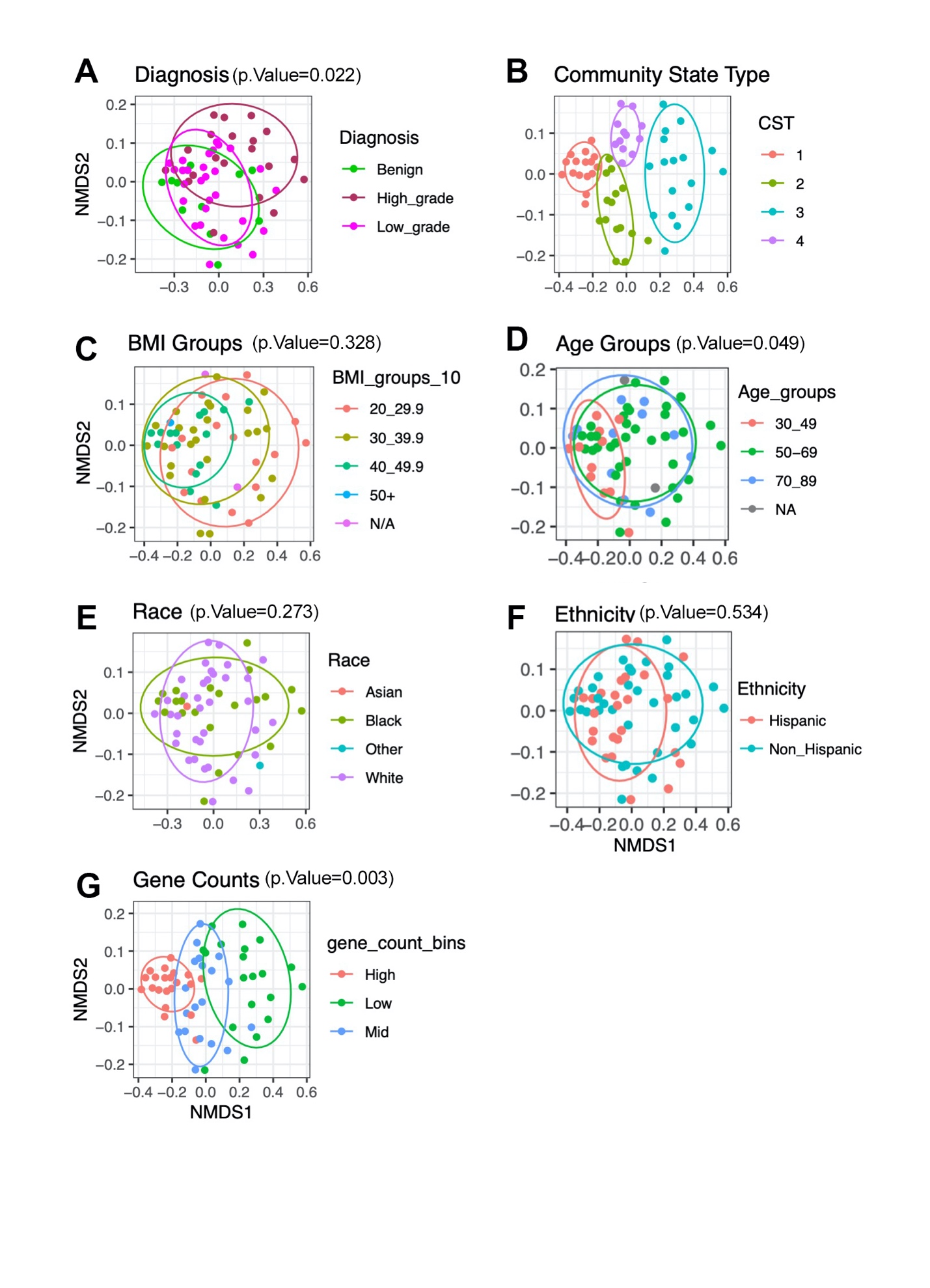

**Supplement 4**. PCA visualization of beta diversity based on seven different clinical variables.
