## Supplement 5 for "Vaginal microbiome is associated with endometrial cancer grade and histology"

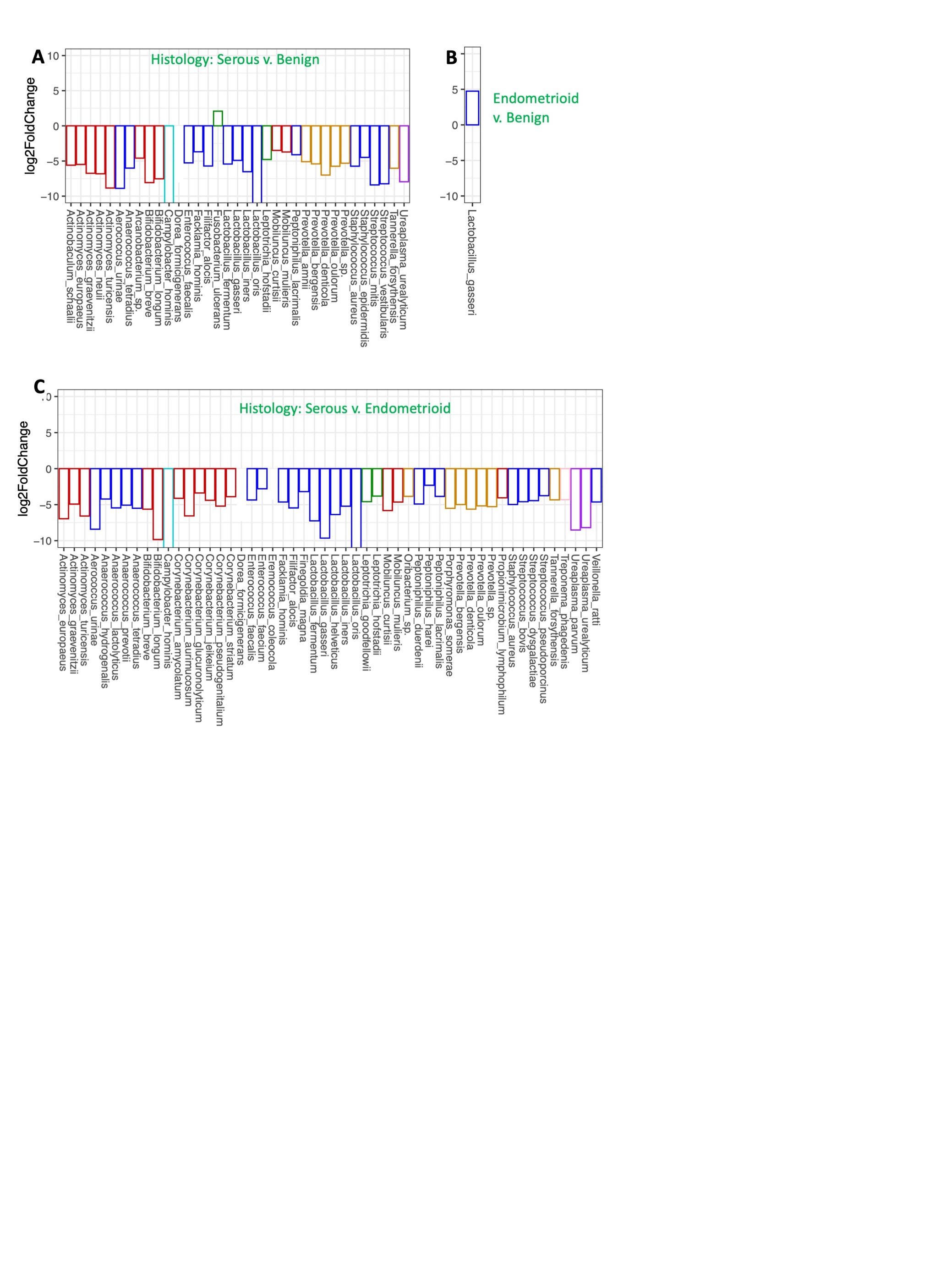

**Supplement 5.** Microbial abundance by histotype. Only taxa with significant changes in abundance are shown in the figure (adjusted p.value<0.05, Wald test). **A**. Serous versus benign. **B**. Endometrioid vs benign. **C**. Serous versus endometrioid.
