## Supplement 6 for "Vaginal microbiome is associated with endometrial cancer grade and histology"

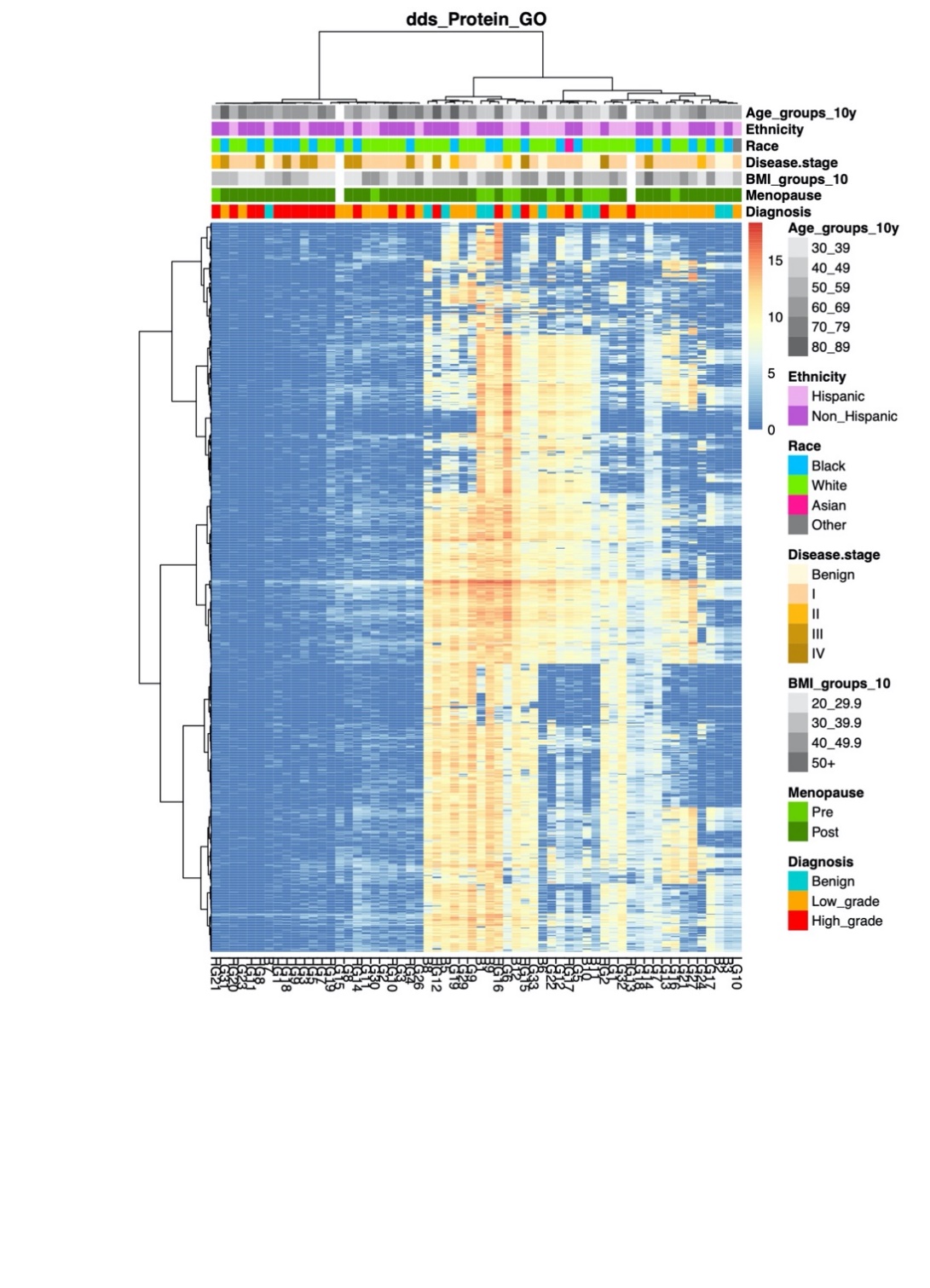

**Supplement 6.** Heatmap of protein functions enrichment score for 61 vaginal metagenomes (red: high enrichment, blue: low enrichment). Functional categories were defined using protein Gene Ontology (proteinGO). Hierarchical clustering of the profiles was performed using ward linkage based on their Euclidean distance.
