## Supplement 7 for "Vaginal microbiome is associated with endometrial cancer grade and histology"

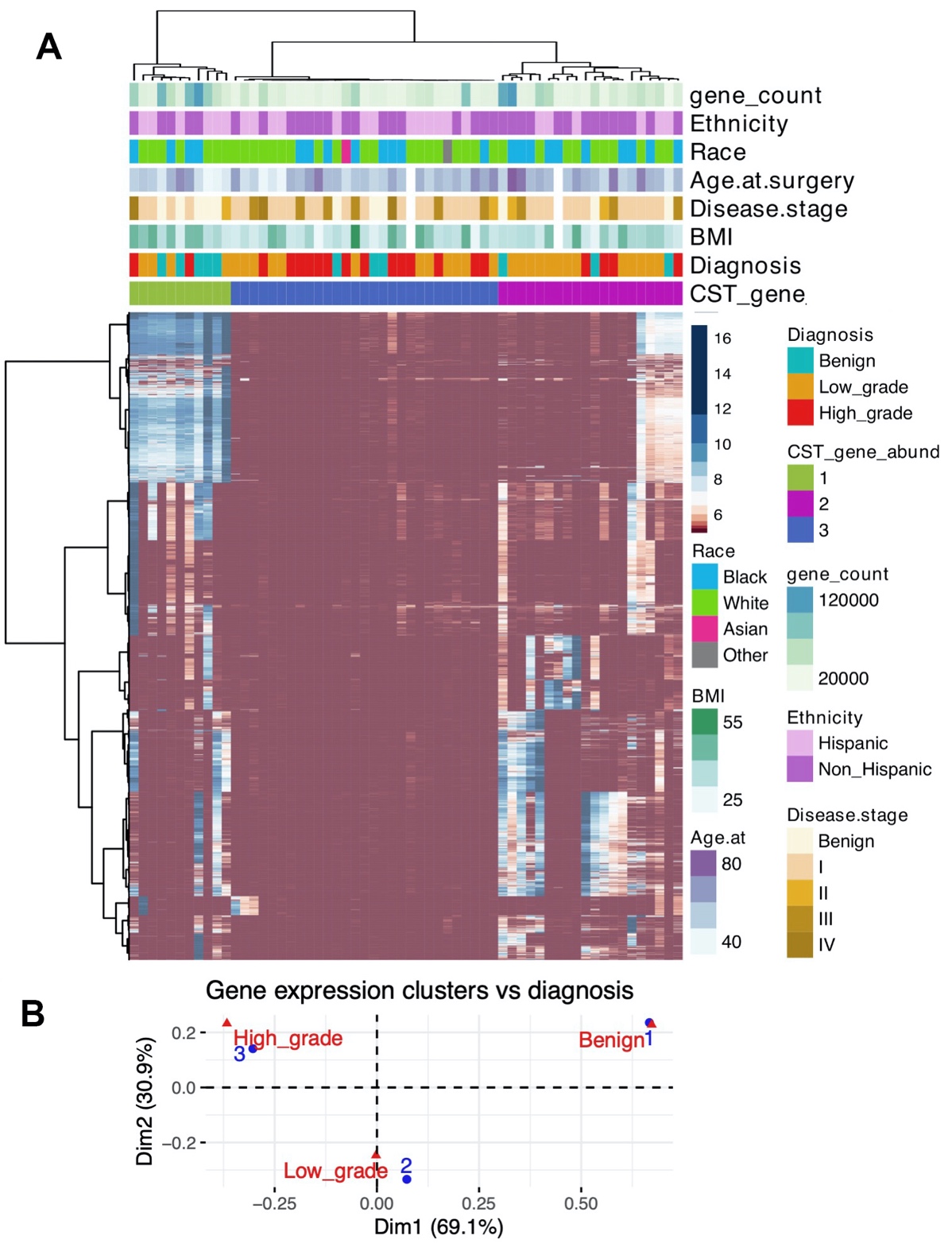

**Supplement 7. A**. Heatmap of abundance values of non-redundant gene profiles for 61 vaginal metagenomes (red: low gene count value, blue: high count value). Hierarchical clustering of the profiles was performed using ward linkage based on their Euclidean distance. **B**. Biplot illustrates the result of correspondence analysis conducted for gene richness and diagnosis. Low grade tumors are more frequent in high gene count communities, while high grade tumors are more frequent in low gene count communities.
