## Supplement 8 for "Vaginal microbiome is associated with endometrial cancer grade and histology"

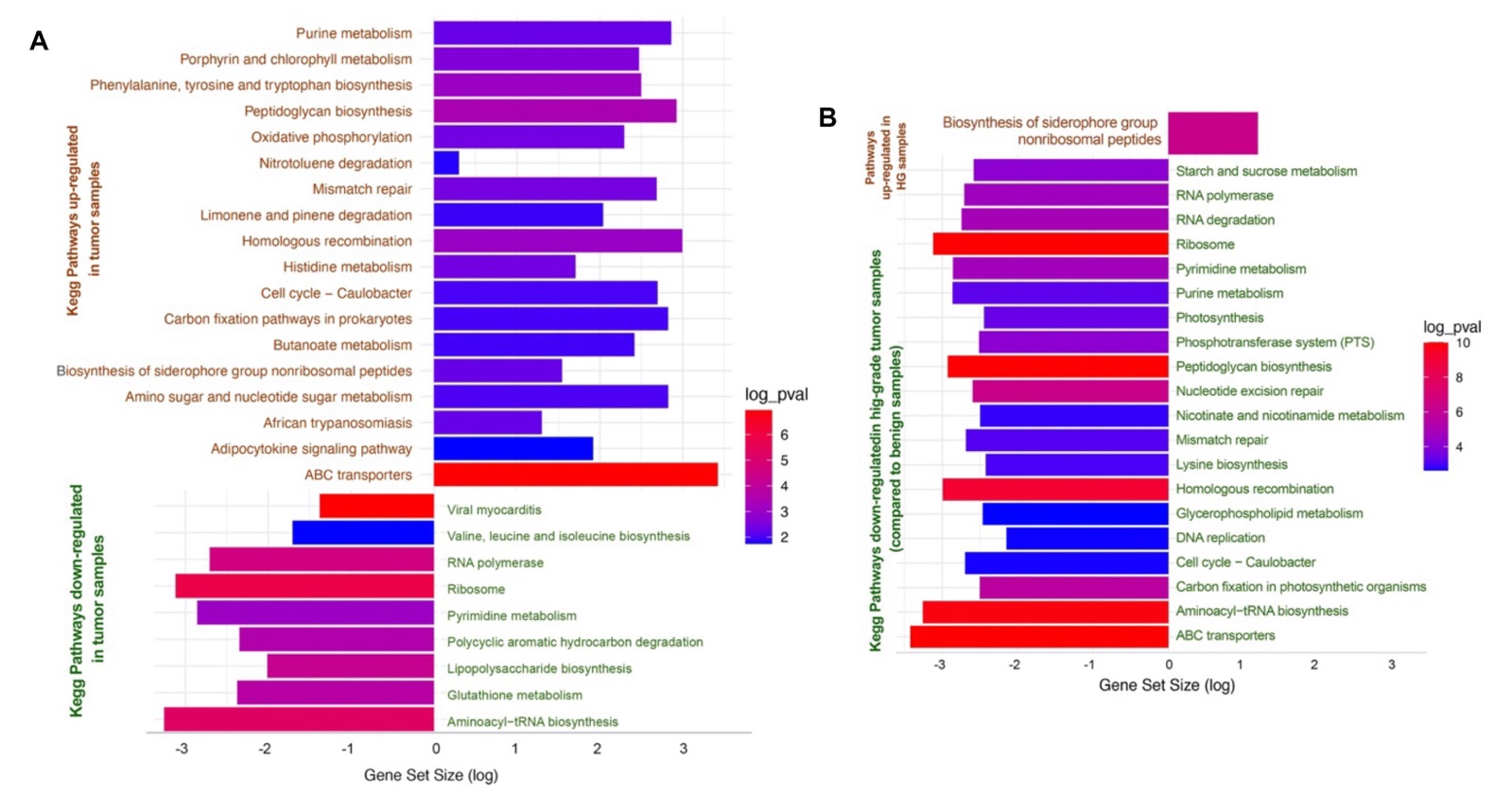

**Supplement 8.** Functional analysis represented by KEGG pathways. **A.** Pathways represented by gene abundance in the metagenomes of endometrial tumors (HG + LG) compared to benign (up or down). **B.** Pathways represented by gene abundance in the metagenomes of HG-endometrial tumors alone compared to benign (up or down). All shown pathways are statistically significant (p<0.05).
