## Supplement 9 for "Vaginal microbiome is associated with endometrial cancer grade and histology"

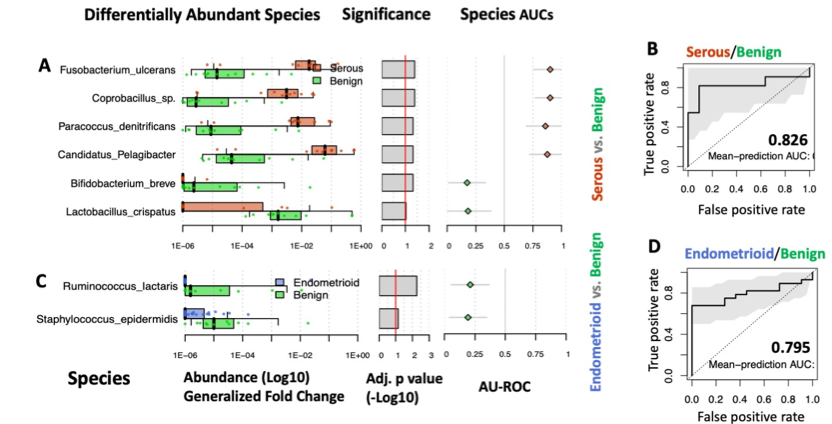

**Supplement 9. Biomarker Discovery by histology.** Additional validation was performed on random forest classifier models, which identified an optimal microbiome signature for each cohort (A, C). These signatures were used to construct receiver operating curves which discriminate serous histology and benign (B), and endometrioid histology versus benign (D).
